# Brain-heart interactions are associated with mortality and acute encephalopathy in ICU patients with severe COVID-19

**DOI:** 10.1101/2024.10.01.24314706

**Authors:** Bertrand Hermann, Sarah Benghanem, Estelle Pruvost-Robieux, Tarek Sharshar, Martine Gavaret, Alain Cariou, Jean-Luc Diehl, Diego Candia-Rivera

**Affiliations:** Université Paris Cité, Inserm UMR-S 1266, Institute of Psychiatry and Neurosciences of Paris, F-75014 Paris, France; GHU Paris Psychiatrie Neurosciences, Service hospitalo-universitaire de Neuro-anesthésie réanimation, F-75014 Paris, France; Service Médecine Intensive - Réanimation, AP-HP. Centre, Hôpital Européen Georges Pompidou, F-75015 Paris, France; Service Médecine Intensive - Réanimation, AP-HP. Centre, Hôpital Cochin, F-75014 Paris, France; Neurophysiology and Epileptology Department, GHU Paris Psychiatrie et Neurosciences, Paris, France; Paris-Cardiovascular-Research-Center, INSERM U970, Paris, France; Université Paris Cité, INSERM, Innovative Therapies in Haemostasis, Paris, France; Sorbonne Université, Paris Brain Institute (ICM), INRIA, CNRS UMR 722, INSERM U1127, AP-HP Hôpital Pitié-Salpêtrière, Paris, France

**Keywords:** Brain-heart interaction, autonomic dysfunction, heart rate variability, COVID-19, acute encephalopathy

## Abstract

**Objective:** Research in critical care has revealed the significance of autonomic dysfunctions, and more recently of brain-heart interactions, as valuable biomarkers for evaluating patients’ physiological status. These biomarkers provide insights into consciousness levels, severity, and outcomes. This study aims to determine the potential of these biomarkers in predicting the mortality and neurological outcome of severe COVID-19 patients.

**Methods:** We examined severe COVID-19 patients who required mechanical ventilation and observed them both during sedation and after sedation cessation. Standard EEG and ECG recordings were conducted at bedside, from which 5 minutes of continuous data were analyzed. Using a synthetic data generation model, we evaluated bidirectional brain-heart interactions from EEG power and heartbeat dynamics series.

**Results:** Our findings indicate that brain-heart interactions, especially involving cardiac parasympathetic activity, can provide information about patients’ severity. We observed correlations with acute encephalopathy duration (coma and delirium), particularly evident in top-down markers (from brain to heart) while bottom-up signaling (from heart to brain) correlated with ICU mortality. Additionally, we noted stronger modulation of brain-heart interactions in milder patients when comparing sedation versus non-sedation conditions, compared to those in more severe states.

**Conclusions:** Our results imply that autonomic dysfunctions, as measured through brain-heart interactions, can track the pathophysiology of comatose states following COVID-19 infection.

**Significance:** These findings highlight the potential of brain-heart interactions as an integrated marker for autonomic function in critical care, offering a more accurate assessment of patient severity and outcomes compared to isolated cardiac or brain measures.

**Highlights:**

- Brain-heart interactions provide valuable insights into patient severity and neurological outcomes in severe COVID-19 pneumonia.
- Brain-to-heart markers correlate with acute encephalopathy duration, while heart-to-brain signaling predicts ICU mortality.
- Brain-heart interactions are modulated differently based on assessment timing and the presence of acute encephalopathy.

## Introduction

In the landscape of COVID-19, the spectrum of neurological manifestations in hospitalized patients ranges from mild headaches to severe presentations such as seizures, encephalitis or acute demyelinating neuropathies (Mao et al., 2020). The underlying mechanisms behind these neurological manifestations remain poorly understood. Among the numerous neurological complications in COVID-19 patients, non-specific acute encephalopathy with prolonged coma and delirium emerges as a significant concern. Indeed, non-specific systemic insults, including inflammation, vasculopathy, and hypoxemia are common and further complicate the treatments (Newcombe et al., 2021; Scott-Solomon et al., 2021). Exploring potential risk factors for acute encephalopathy reveals insights into the evolving pathology, where the transition from mild to severe often encompasses stages of delirium progressing towards coma (Helms et al., 2020; Pun et al., 2021).

Central to our understanding of severe COVID-19 manifestations is the role of the brainstem, serving as a critical hub for motor function, sensory processing, arousal, respiratory control, autonomic regulation, and neuroimmune mechanisms (Benghanem et al., 2020). Brainstem reflexes, while non-specific, hold high prognostic value, making them valuable tools in clinical assessment (Kandelman et al., 2020; Rohaut et al., 2017; Sharshar et al., 2011). The regulatory function linked to the autonomic nervous system suggests a potential connection with the pathophysiology of COVID-19. The dysfunctions in autonomic mechanisms appear to be a significant feature in many critical care conditions, notably in sepsis (Carrara et al., 2021). Recently, we showed that brain-heart interactions in post-cardiac arrest hypoxic ischemic encephalopathy signal the severity and prognosis (Hermann et al., 2024). While most studies have primarily focused on assessing cardiac autonomic dysfunction through measures of heart rate variability (HRV) (Benghanem et al., 2024; Endoh et al., 2019), the assessment of bidirectional brain-heart interactions proved useful in further characterizing the patients. Indeed, these findings are in line with recent efforts to stratify post-comatose patients based on their wakefulness and responsiveness (Candia-Rivera and Machado, 2023), in which a preserved brain-heart communication and its variability were indicative of the presence and level of consciousness.

Considering this evidence, our present study investigates the relationship between COVID-19 disease complications and brain-heart interactions. We focused on adults admitted in the intensive care unit (ICU) for severe COVID with acute respiratory distress syndrome (ARDS) requiring invasive mechanical ventilation and deep sedation. Our investigation aims to investigate how brain-heart interactions can elucidate the physiological state from an early stage. We examined EEG and ECG recordings from patients within the first 12-72 hours following neuromuscular blocking agent cessation (T1), and between 3 to 7 days after sedation weaning (T2). We aimed at contrasting the brain-heart interactions at these two timepoints, as a function of the patient complications, and to investigate if early brain-heart interactions were associated with ICU outcome (survival and acute encephalopathy-free days at day 28). To achieve this, we conducted an analysis of brain-heart interactions by modeling interactions between EEG and cardiac time series (Candia-Rivera, 2023). This study seeks to explore two key questions regarding brain-heart interactions in severe COVID-19 ICU patients. First, it aims to examine whether early brain-heart interactions, potentially indicative of dysautonomia, hold prognostic value for patient outcomes. Second, it investigates how these interactions evolve during the ICU stay and potentially track the neurological/behavioral status of patients.

## Materials and Methods

### Population

This was an ancillary analysis of the Brainstem-CoV-ICU study (Benghanem et al., 2022). Briefly, the Brainstem-CoV-ICU study was a bicentric prospective observational study investigating the prevalence and prognostic significance of an early clinical and standard EEG assessment in ICU COVID-19 patients. Adult patients included were admitted in the ICU for a severe, proven SARS-CoV-2 pneumonia, leading to an acute respiratory distress syndrome (ARDS) and requiring invasive mechanical ventilation with initial deep sedation (Richmond Agitation-Sedation Scale <-3 for at least 12h). Patients with previous central neurologic disorders were excluded.

IRB approval was granted by *Comité de Protection des Personnes ‘Ile-de-France II’* (n°ID RCB: 2020-01559-30) and the study was a priori registered on clinicaltrials.gouv: NCT04527198. A written informed consent was obtained from each of the patients or their relatives.

### Design and data collection

Detailed neurological assessments and EEG recordings were performed at two timepoints during the ICU stay (Figure 1A). The first (T1) was under sedation, 12-72h following neuromuscular blocking agents’ withdrawal if any, and the second (T2) was 3-7 days after definite sedation withdrawal. At each timepoint, concomitant organ failures (based on the Sequential Organ Failure Assessment score, SOFA) and sedative/opioid exposure (duration, infusion rates and cumulative doses at time of assessments) were collected.

**Figure 1.**
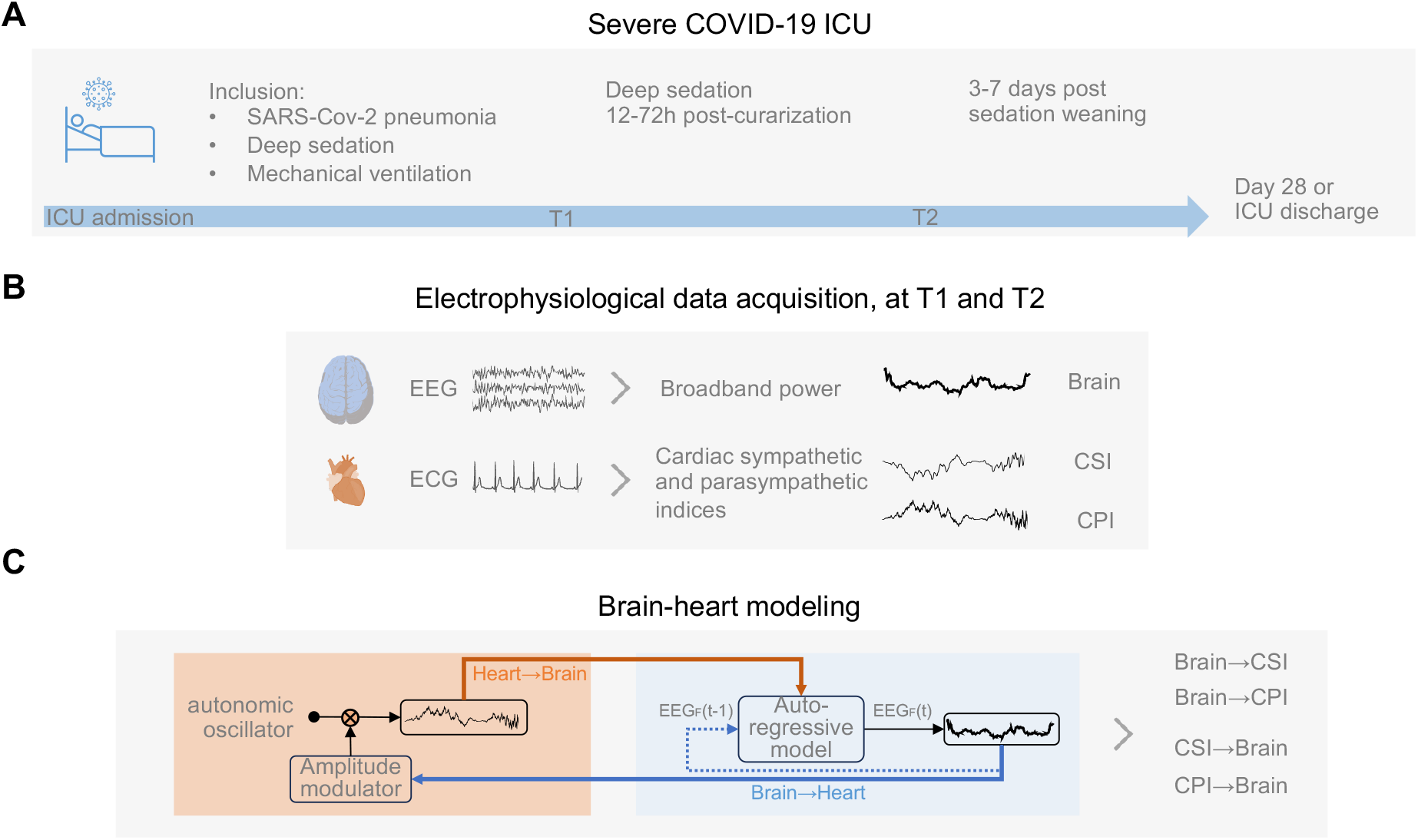
Computation of brain-heart interactions through a synthetic data generation model. (A) Severe COVID-19 patients admitted in the ICU. (B) Data acquisition was performed under sedation, 12-72h following neuromuscular blocking agents’ withdrawal (T1) and 3-7 following sedation weaning (T2). (C) Bidirectional brain–heart interactions between the brain within the band of interest (1-12 Hz) and cardiac sympathetic (CSI) and parasympathetic indices (CPI). The model estimates the functional interplay between the brain and the heart by assuming a closed-loop circuit, in which ongoing EEG power modulates autonomic activity, while in turn, ongoing autonomic activity (CSI and CPI) modulates EEG activity.

Neurological status was assessed by the Richmond Agitation-Sedation Scale (RASS) every 4h and by the Confusion Assessment Method for the ICU (CAM-ICU) twice daily during the 28-days following T1 or up to ICU discharge, whichever came first. Acute encephalopathy was defined as a state of coma (RASS<-3) or delirium (positive CAM-ICU in patients with RASS≥-3). Additionally, we defined awakening as two consecutive RASS≥-2 and delayed awakening as the absence of awakening 3 days after sedation discontinuation (Pun et al., 2021). Last, vital status at 28 days and at ICU discharge was also collected.

### EEG/ECG recordings and preprocessing

Standard video-EEG recordings were acquired with 11 scalp electrodes (10-20 international system: Fp1, C3, T3, O1, Fp2, C4, T4, O2, Fz, Cz and Pz) and 1 ECG lead for 20 minutes. Impedances were kept below 5 kΩ. Sampling rate was 256 Hz.

The ECG signals were filtered using a band-pass filter from 0.5 to 20Hz. EEG signals were filtered using a band-pass filter from 0.5 to 45Hz. Butterworth filters of order 6 and 8 were applied for ECG and EEG filtering, respectively. Notch filters were employed at 50Hz and 100Hz, and the signals were re-referenced to an average reference. Independent component analysis (ICA) was then applied to the EEG signals, following a high pass filtering at 1Hz. The resulting components were visually examined for significant artifacts, such as eye movements, blinks, or cardiac-field artifacts. The artifact-free EEG was then compared to the original recording to evaluate the quality of artifact removal and preservation of brain EEG signals. Subsequently, the reconstructed recordings underwent visual inspection, and periods containing residual artifacts were manually excluded. The initial 5-minute period of consecutive data without EEG and ECG artifacts was retained for further analysis.

EEG power spectrum densities were computed using a short-time Fourier transform with a Hamming taper using a sliding window procedure (2 second segments with 50% overlap). Based on our previous findings, we integrated the EEG power series within 1-12Hz (Hermann et al., 2024). In case of residual noise, individual EEG channels were denoised using Wavelet thresholding method (Jansen, 2001), with a threshold defined automatically, based on signal’s length and variance (Gabard-Durnam et al., 2018).

### Brain-heart interaction modeling

Bidirectional brain-heart interactions were computed using a synthetic data generation model (Candia-Rivera, 2023), between EEG power series within the 1-12 Hz band, hereafter called broadband power, and cardiac sympathetic (CSI) and parasympathetic indices (CPI). The synthetic data generation framework quantifies time-resolved relationships between ongoing fluctuations in brain and cardiac time series. On the heart side, the framework aims to model the stimulations to the sinoatrial node that causes the heartbeat generation. On the brain side, the framework models the fluctuations on EEG power on time as an autoregressive process, with an external input (CSI or CPI). The outputs of the model are time-varying coefficients accounting for the brain-heart interactions for each combination of EEG power series, CSI or CPI, and ascending or descending directionality, as depicted in Figure 1B and C. The computation of brain-heart interplay coefficients was done over EEG power series (mV^2^) and CSI and CPI series (arbitrary units) resampled at 1 Hz, using a 15 s sliding time window. To quantify the overall strength of the brain-heart coupling, absolute values of the resulting time-varying coefficients were averaged among channels and on time using the median over the 5 minutes time series and log-transformed (arbitrary units).

### Exposures and outcomes

Main exposures were the median brain-heart interplay coefficients from and towards both cardiac sympathetic and parasympathetic indices, that is four metrics in total: brain→CSI, brain→CPI, CSI→brain and CPI→brain.

The two main outcomes were ICU survival and the burden of acute encephalopathy, expressed as the time to recovery from the acute encephalopathy, within the 28 days from T1 (days-free of coma and delirium for at least 48h). Other outcomes were delayed awakening and neurological status at T2. For further analyses, brain-heart coupling values were log10 scaled.

### Statistical analyses

Continuous variables were expressed as medians and interquartile ranges. Paired and unpaired comparisons were performed using Wilcoxon-Mann-Whitney tests. Qualitative variables were expressed as number and percentages and compared using chi-square or Fisher test as appropriate.

To assess the multiple variables associated with the patients and their potential interactions, we used multiple linear regression models of the logarithm of the hazard on the variables (multivariable Cox). This allowed us to investigate the association of brain-heart interplay metrics and both the cumulative incidence of survival and of acute encephalopathy recovery at day 28 from T1. Besides the four brain-heart metrics, univariable models were built with non-neurological SOFA, RASS score, sedative/analgesic instantaneous rate, cumulative dose (in midazolam equivalent, assuming that 10mg of propofol would be equal to 1 mg of midazolam) and duration at T1. P-values were computed with the likelihood ratio test. An exploratory multivariable analysis with adjustment on variables with p-values<0.2 on univariable analysis was performed. Additionally, as sedation might be associated with both mortality and encephalopathy, and influence brain-heart interactions, multivariable models adjusted on sedation-related variables were performed.

Lastly, evolution of brain-heart metrics over time (from T1 to T2) according to neurological status at T1 was modeled with aligned rank transformed for non-parametric factorial analysis of variance. Mixed ANOVA with the timing as within-subject factor and neurological status as between-subject factor was performed on aligned rank transformed data (package ARTool).

All tests were two-sided with p-values of less than 0.05 considered as significant. Statistical analyses were performed using R Software, R version 4.3.3 (2024-02-29); https://cran.r-project.org/).

## Results

### Population

From April to December 2020, out of the 52 patients included in the Brainstem-CoV-ICU study, 49 could be included in this study (3 patients were discarded due to less than 5 minutes of electrophysiological recordings of sufficient quality). Patients were mostly men (82%) aged 68 [56-73] years. Median SAPS2 and SOFA at ICU admission were 48 [35-67] and 5 [4-10]. ARDS severity was moderate in 55% and severe in 45% with 88% of patients treated with dexamethasone. Population characteristics are presented in Table 1.

**Table 1.** Population and ICU outcome.

| Characteristic | N = 49 |
| --- | --- |
| **Age (years)** | 68 [56-73] |
| **Female sex** | 9 (18) |
| **BMI (kg/m<sup>2</sup>)** | 27.3 [25.0-29.6] |
| **Obesity (BMI ≥30 kg/m<sup>2</sup>)** | 13 (27) |
| **MacCabe score** |  |
| 1 | 39 (80) |
| 2 | 10 (20) |
| **SAPS2 at ICU admission** | 48 [35-67] |
| **SOFA at ICU admission** | 5 [4-10] |
| **ARDS severity** |  |
| Moderate ($100 < \text{PaO}_2/\text{FiO}_2 \leq 200$) | 27 (55) |
| Severe ($\text{PaO}_2/\text{FiO}_2 \leq 100$) | 22 (45) |
| **Minimal PaO<sub>2</sub>/FiO<sub>2</sub>** | 106 [85-119] |
| **Awakening** |  |
| Yes | 40 (82) |
| Delay (days) | 3 [0-11] |
| Delayed | 26 (65) |
| **Acute encephalopathy duration (days)** | 16 [11-29] |
| Coma incidence (%) | 49 (100) |
| Coma duration (days) | 12 [7-25] |
| Delirium incidence (%) | 30 (75) |
| Delirium duration (days) | 5 [3-8] |
| Coma/Delirium-free days at day 28 from T1 | 14 [0-25] |
| **ICU mortality** | 12 (24) |

The first assessment was performed in median 4 [3-6] days after ICU admission in comatose patients under deep sedation. Details about neurological and sedative status at T1 are presented in Table 2. There were no significant correlations between any of the brain-heart metrics and either non-neurological SOFA score or the RASS score.

**Table 2.**
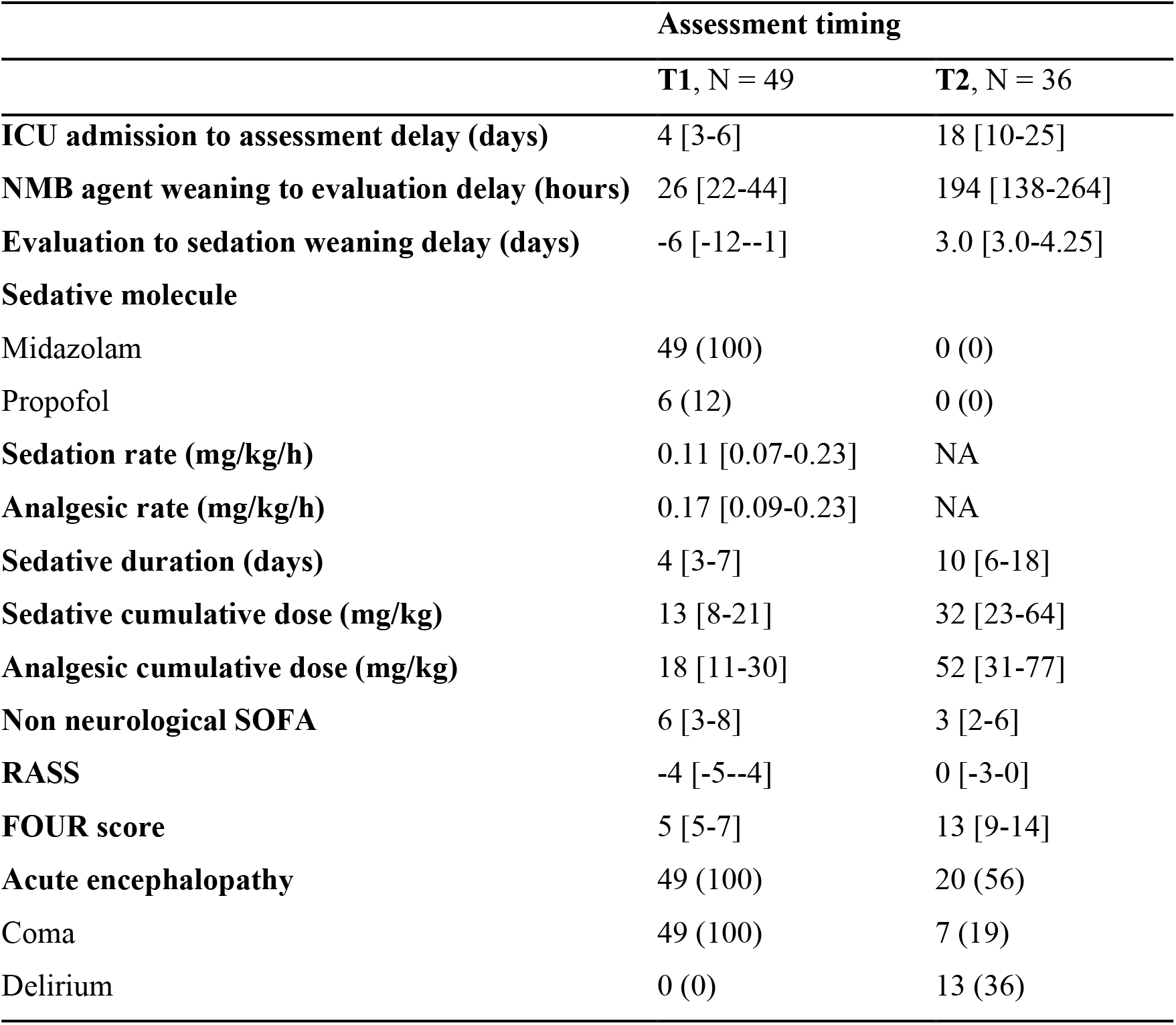
Neurological and sedative status at T1 and T2 assessments.

### Early heart→brain interactions are associated with mortality

Twelve patients (24%) died during their ICU stay. All brain-heart interactions metrics measured at T1 significantly differed between patients who died vs. those who survived. Death was associated with stronger brain-to-heart coupling and lower heart-to-brain coupling (Figure 2).

**Figure 2.**
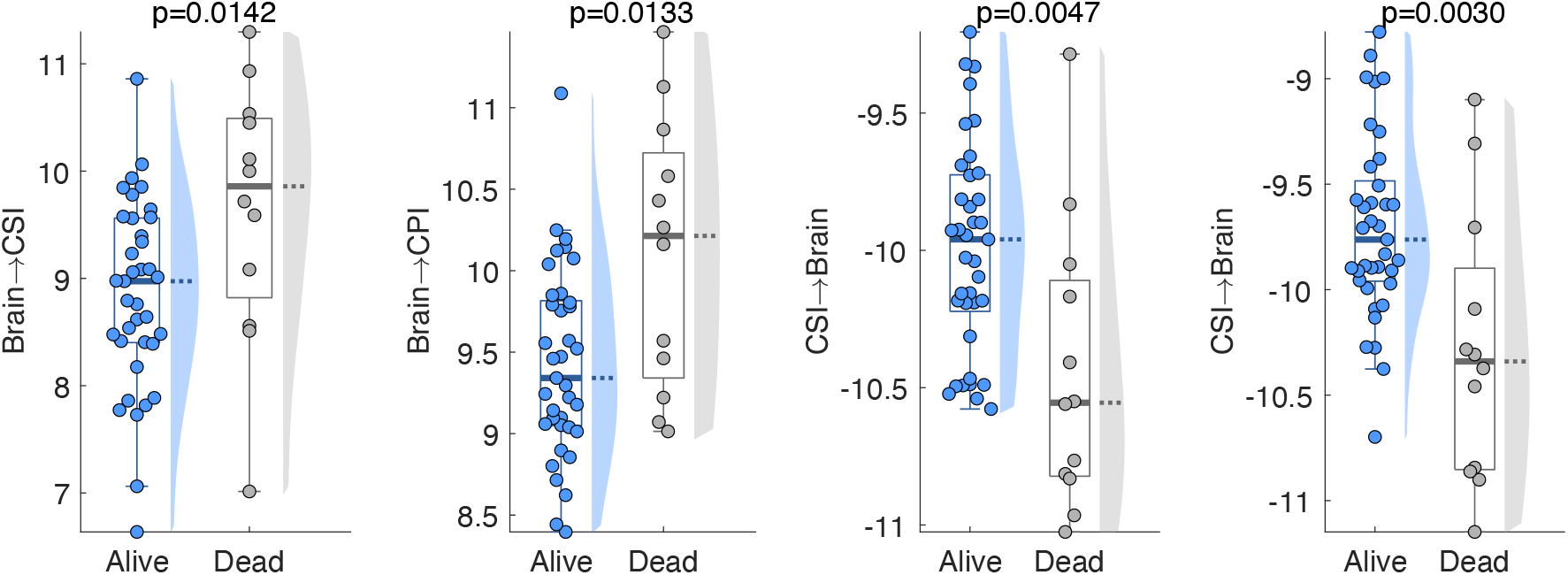
Brain-heart metrics according to vital status at ICU discharge. P-values correspond to the unpaired Wilcoxon test, with significance defined by the Bonferroni rule for multiple comparisons at α=0.0125. Brain: EEG power integrated within 1-12 Hz, CSI: cardiac sympathetic index, CPI: cardiac parasympathetic index.

To refine this association, we ran time-to-event analyses investigating the association of brain-heart metrics with the cumulative incidence of death during the 28 days from T1 follow-up using Cox models. In univariable analyses, both heart→brain metrics, CSI→brain and CPI→brain, were significantly associated with the cumulative incidence of death (crude HR 0.21 [0.05-0.85], p=0.022 and HR=0.24 [0.07-0.77], p=0.016 respectively) (Supplementary table 1). On multivariable analyses CPI→brain remained independently associated with mortality in all models, with lower coupling values being associated with higher cumulative incidence of death (Table 3). This effect seemed also independent of sedation regimen as the association persisted after adjustment on either instantaneous rate, cumulative dose or duration of sedative and analgesic at T1 (Supplementary table 2).

**Table 3.** Cox regression models for associations of brain-heart metrics with mortality and encephalopathy at day 28. Univariable and multivariable Cox regressions models. In multivariable analyses, hazard ratios were adjusted on variables with p-values<0.2 on univariable analysis, that is RASS, non-neurological SOFA and sedative rate for mortality and RASS and sedative rate for encephalopathy.

| Mortality at day 28 from T1 |  |  |  |  |
| --- | --- | --- | --- | --- |
| Univariable |  |  | Multivariable |  |
| Independent Variables | HR [95% CI] | p | aHR [95% CI] | p |
| CSI → Brain | 0.21 [0.05-0.85] | **0.022** | 0.24 [0.06-1.06] | 0.060 |
| CPI → Brain | 0.24 [0.07-0.77] | **0.016** | 0.24 [0.07-0.82] | **0.022** |
| Recovery from coma and delirium at day 28 from T1 |  |  |  |  |
| Univariable |  |  | Multivariable |  |
| Independent Variables | HR [95% CI] | p | aHR [95% CI] | p |
| Brain → CPI | 0.38 [0.18-0.8] | **0.005** | 0.43 [0.2-0.95] | **0.036** |
| CSI → Brain | 2.76 [1.18-6.42] | **0.016** | 2.35 [0.95-5.81] | 0.064 |
| **CPI → Brain** | 2.32 [1.09-4.93] | **0.025** | 1.78 [0.8-3.96] | 0.155 |

### Early brain→heart interactions are also associated with acute encephalopathy

Median acute encephalopathy duration was 16 [12-29] days, with median duration of coma of 12 [7-25] and a median duration of delirium of 5 [3-8] days in the 30 (75%) patients that developed delirium (Table 2).

As for the mortality outcome, we assessed the association of brain-heart metrics with coma-and delirium-free days, using Cox regression models. In univariable analyses, both heart→brain metrics were significantly associated with the cumulative incidence of acute encephalopathy recovery within 28 days from T1 (HR 2.76 [1.18-6.42], p=0.016 for CSI→brain and HR 2.32 [1.09-4.93], p=0.025 for CPI→brain), as well as brain→CPI (HR=0.38 [0.18-0.80]) (Supplementary table 1). In multivariable analyses, only brain→CPI remained significantly associated with acute encephalopathy, with higher coupling being associated with lower coma/delirium-free days at day 28 (Table 3). Again, this association was independent of sedative/analgesic (Supplementary table 2).

Lastly, 9 patients died before sedation could be stopped and delayed awakening was observed in 26 (65%) of the remaining 40 patients. Brain→CPI at T1 was also significantly higher in patients with delayed awakening (9.66 [9.31-9.99] vs. 9.05 [8.87-9.10], p=0.001).

### Brain-heart metrics outperform individual brain and heart metrics

Finally, we examined whether heart and brain metrics alone could explain some of the relationships found in brain-heart interactions. EEG power was significantly associated with ICU mortality (AUC=0.75 [0.56-0.93]) but with lower AUC than heart-to-brain metrics (CPI-to-brain AUC=0.79 [0.6-0.98] and CSI-to-brain AUC=0.77 [0.59-0.96]). CSI and CPI metrics derived from the ECG were not associated with ICU mortality (Figure 3A, Supplementary table 3). EEG power was also associated with delayed awakening (AUC=0.69 [0.51-0.87]), but again with a lower AUC than brain-to-CPI (AUC=0.8 [0.63-0.97]). Again, CPI and CSI were not associated with delayed awakening (Figure 3B, Supplementary table 4). Lastly, a significant correlation with acute encephalopathy duration was only found with brain-to-CPI (rho=0.44, p=0.004) while CSI, CPI and EEG power were not (Figure 3C).

**Figure 3.**
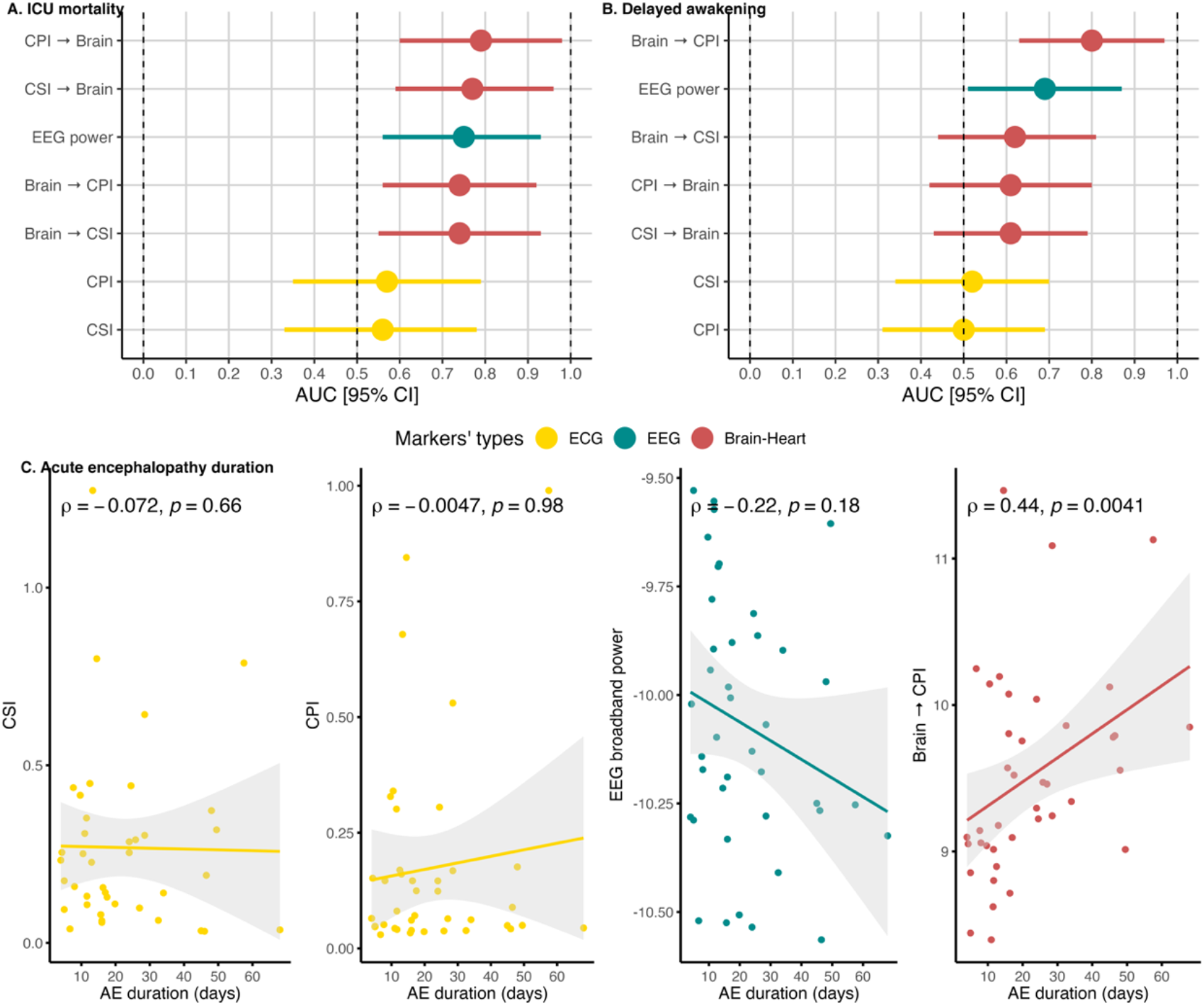
Relationships between ECG metrics (CSI and CPI, yellow), EEG metric (EEG power, green) and brain-heart metrics (red) at T1 and different outcomes. (A) ICU mortality, (B) delayed awakening and (C) acute encephalopathy (AE) duration. For ICU mortality and delayed awakening, area under the ROC curve (AUC) and corresponding 95% confidence interval (95% CI) are represented. Correlation with AE duration was computed using spearman correlation coefficients (rho).

### Acute encephalopathy modulates the evolution of brain-heart interactions

At T2, that is 3-7 days after sedative withdrawal, 36 (73%) patients could be assessed, 13 were excluded because 7 died before, EEG was not performed in 3 of them, and insufficient data quality in other 3 of them. Regarding neurological status, 20 (56%) patients still suffered from acute encephalopathy, being either comatose (19%) or delirious (36%) (Table 2). We then investigated the changes over time (from T1 to T2, within-subject factor) according to neurological status at T2 (acute encephalopathy vs. non-acute-encephalopathy, between-subject) using an aligned rank transformed mixed ANOVA (from T1 to T2).

For brain→heart metrics, only a significant main effect of timing was found (F(1, 31)=14.6, p<0.001 for brain→CSI, F(1,31)= 20.9, p<0.001 for brain→CPI), with increasing coupling values over time. While in the opposite direction, a significant timing × neurological status interaction was found for both CSI→brain (F(1, 31)=8.1, p=0.008) and CPI→brain (F(1, 31)=7.6, p=0.01). Post-hoc comparisons showed that a significant reduction of heart-to-brain coupling from T1 to T2 was only observed in patients without acute encephalopathy at T2 (T1 vs. T2: CSI→brain, p=0.0001; CPI→brain, p=0.0003), as compared to patients with acute encephalopathy (CSI→brain, p=0.3512; CPI→brain, p=0.6152), as shown in Table 4 and Figure 4.

**Figure 4.**
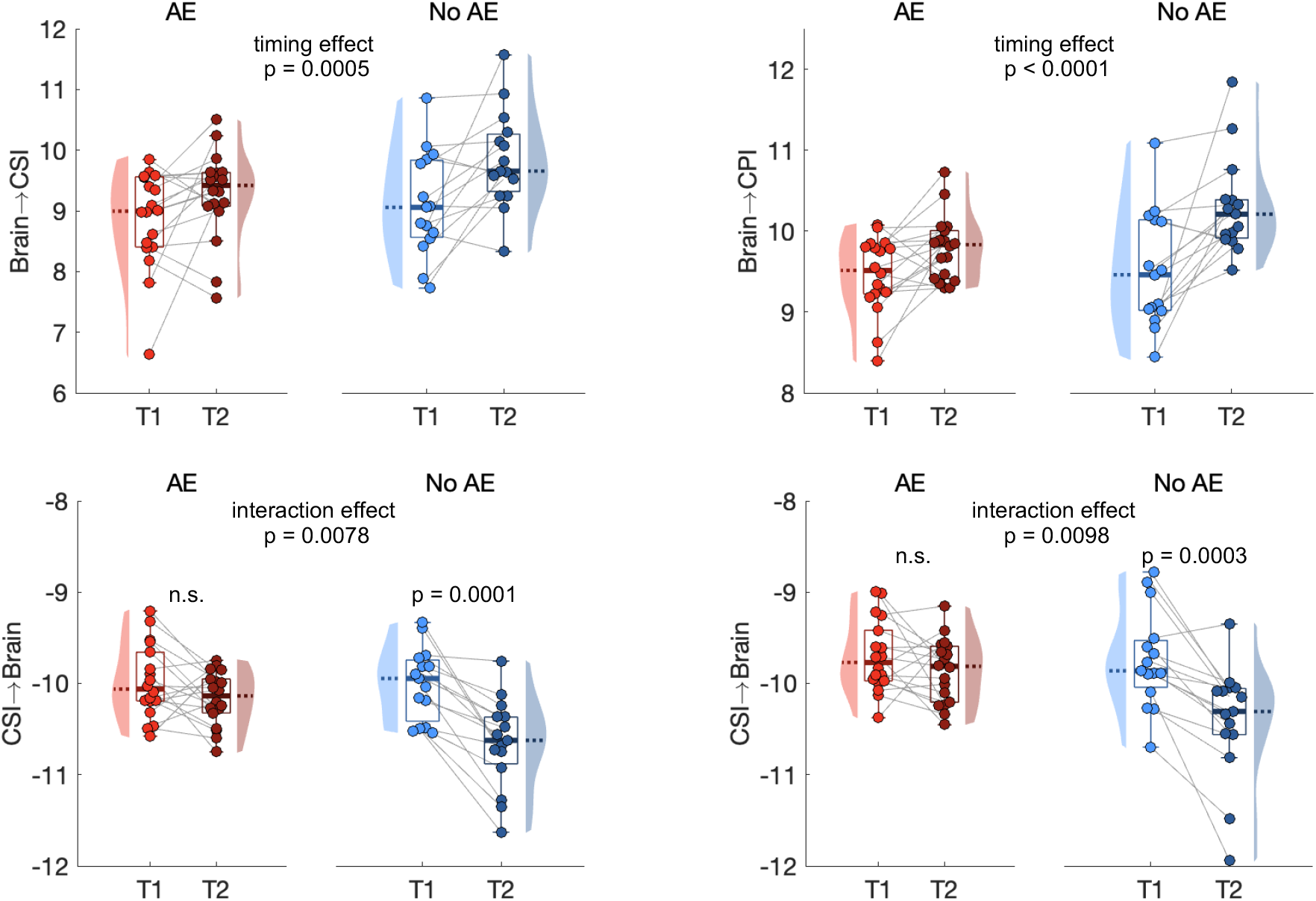
Evolution of brain-heart metrics (from T1 to T2) according to neurological status at T2 (with or without acute encephalopathy). P-values correspond to the ANOVA analysis with respect to the timing (T1 or T2), state at T2 (AE or no AE), and their interaction. CSI: cardiac sympathetic index, CPI: cardiac parasympathetic index. AE: acute encephalopathy.

**Table 4.** ANOVA analysis.

| Effect | Timing | | State (at T2) | | Interaction: Timing × State | |
| --- | --- | --- | --- | --- | --- | --- |
| values | F(1,31) | p | F(1,31) | p | F(1,31) | p |
| brain→CSI | 14.6359 | **0.0006** | 1.8907 | 0.1790 | 1.2887 | 0.2650 |
| brain→CPI | 20.9259 | **<0.0001** | 1.7288 | 0.1982 | 3.9497 | 0.0558 |
| CSI→brain | 21.8315 | **<0.0001** | 4.2738 | **0.0471** | 8.1019 | **0.0078** |
| CPI→brain | 18.6688 | **0.0001** | 3.9892 | 0.0546 | 7.5864 | **0.0097** |

## Discussion

The autonomic nervous system plays a crucial role in mediating the effects of brain complications on the heart, involving neural, endocrine, and immune mechanisms, such as through the hypothalamus-pituitary-adrenal axis (Chen et al., 2017). While the multisystem alterations occurring in severe COVID-19 pneumonia are acknowledged complications (Newcombe et al., 2021; Scott-Solomon et al., 2021), these multisystem manifestations remain poorly understood. In our prospective study, we investigated brain-heart interactions within a cohort of patients hospitalized in the ICU due to severe COVID-19 pneumonia. We uncovered compelling insights into the relationship between these interactions and both patient neurological status and prognosis. Notably, our findings revealed a significant association between brain-heart interactions within the first days of admission to ICU and mortality, and the burden of acute encephalopathy at day 28.

### Brain-heart interactions inform multisystem dysfunctions

We observed that bidirectional brain-heart interactions related to patient’s outcomes, where more severe patients were associated with aberrant interactions: a higher brain→heart (hypercoupling) and lower heart→brain interactions. These results suggest that severe COVID-19 pneumonia leads to systemic changes, including dysautonomia, causing the heart to respond abnormally to most brain changes, but also resulting in reduced central integration of interoceptive signals. Considering that the cardiovascular system maintains a closed-loop communication with cortical areas, including structures such as the insular cortex, anterior cingulate gyrus and brainstem autonomic centers (Nagai et al., 2010), their impairment can lead to changes in specific cardiac rhythms, blood pressure, disrupted levels of brain natriuretic peptide, catecholamines, glycemia and breathing patterns. The latter could contribute to respiratory and weaning failure due to maladaptive central respiratory drive (Esnault et al., 2020; González-Duarte and Norcliffe-Kaufmann, 2020; Manganelli et al., 2020). Actually, dysautonomia appear to be a significant feature of many critical care conditions, notably in sepsis (Carrara et al., 2021). Moreover, COVID-19 infection can include direct influences on cardiovascular systems, including dysregulation of ACE2 enzyme, microvascular dysfunction, hypoxemia and cardiotoxicity (Abha Mishra et al., 2023). The infection might disrupt as well neurovascular coupling, altering cerebral blood flow and the subsequent adjustments in brain activity according to perfusion levels, ultimately affecting cellular homeostasis, unbalancing energy supply and demand (Kim et al., 2016).

### Bidirectional interactions differ based on etiology

Our findings on the relationship between aberrant brain-heart interactions and patients’ severity are in line with our previous work, in which we found that similar patterns also indicated poor neurological outcome after cardiac arrest (Hermann et al., 2024). However, in the present study, heart→brain provided stronger associations with mortality, while in cardiac arrest patients it was primarily the brain→heart metrics that showed a stronger relationship with the severity and outcomes. Although these differences might arise from varying patients’ etiologies, specific pathophysiological mechanisms are likely responsible for them. For instance, from differences in the regions affected in both types of brain injury. In cardiac arrest patients, there are extensive cortical lesions caused by the post-anoxic injury, with relatively spared brainstem (Endisch et al., 2020). Instead, infections involve brainstem responses due to neuroinflammation, such as those well-reported in sepsis (Becher et al., 2017; Benghanem et al., 2020; Sharshar et al., 2003), but also in COVID-19 (Matschke et al., 2020; Thakur et al., 2021; Weyhern et al., 2020).

These results would also indicate that both directions of the brain-heart interactions might assess several components of the autonomic nervous system (from brainstem centers to limbic and neocortical higher-order areas involved in autonomic control) which are differentially involved in vital and neurological outcome. Nonetheless, this top-down hypercoupling with reduced bottom-up signaling, seems to be a hallmark of both hypoxic ischemic brain injury and COVID-19 associated encephalopathy. Indeed, in turn, brain→heart interactions were mainly associated with acute encephalopathy.

### Acute cognitive impairments influence the progression of brain-heart interactions

Longitudinal follow-up of patients who survived until sedation weaning showed that brain-heart interactions were able to track patients’ neurological status during the recovery phase, indicative of the presence of acute encephalopathy (coma and delirium). Indeed, patients with absence of acute encephalopathy, indicative of a milder state, presented a stronger reduction of their heart→brain interactions at T2. Non-specific acute encephalopathies with prolonged coma and delirium are common neurological complications in severe COVID-19 infection (Helms et al., 2020; Pun et al., 2021), and could relate to the degree of hypoxemia (Waldrop et al., 2022). While some early qualitative EEG patterns (i.e. discontinuous and nonreactive background) have been linked to mortality and duration of acute encephalopathy (Benghanem et al., 2022), altered brain-heart interactions appears as a key element in COVID-19 pathophysiology fostering the development of severe multiorgan failure and endothelial injury (Lionetti et al., 2021).

While the relationship between the progression of brain-heart interactions and the presence of acute encephalopathy after sedation weaning seems to appear from a strong modulation at the brain level, disparate physiological couplings are typically observed in general anesthesia (Supp et al., 2011). Furthermore, when patients experience deviations from these trends, it may indicate the presence of significant physiological failures (Maschke et al., 2023). Interestingly, while high brain-heart coupling in the early phase was indicative of more severe states, during the recovery, a higher coupling was associated with milder states, illustrating the complex and dynamic nature of the autonomic response to systemic dysfunctions. This shift underscores the evolving interplay between adaptive and maladaptive responses throughout the phases of critical care.

Finally, our results may lead to future research bridging the gap with the reported brain-heart interactions informing about disorders of consciousness (Candia-Rivera et al., 2023, 2021; Candia-Rivera and Machado, 2023; Pérez et al., 2021; Raimondo et al., 2017). Intriguingly, a preserved brain-heart communication and its variability can serve as a marker of presence and level of consciousness, respectively (Candia-Rivera and Machado, 2023). In light of those results, our study underscores the importance of measuring the directionality of altered brain-heart interactions to better inform acute cognitive impairments across different etiologies.

### Predominant dysregulations in the parasympathetic nervous system

We found that the relationship between aberrant brain-heart interactions and the patients’severity was predominant from and towards the parasympathetic system. Sympathetic activity is generally considered a better marker for assessing the severity of a patient’s condition in critical care (Hilz et al., 2019). For instance, elevated sympathetic activity, such as elevated heart rate or blood pressure, is often associated with acute stress responses, such as in cases of trauma or severe infections (Baguley et al., 2006; Shashikumar et al., 2017). On the other hand, parasympathetic activity, may not show such clear and immediate changes in response to severe physiological status (Hilz et al., 2019), although vagal reflexes play a significant role in balancing physiological responses and relaxation, but also in regulating respiratory rate and immune response. Our results indicate that brain-heart interactions, particularly involving parasympathetic activity, reveal clearer differences in patient states and outcomes. This finding suggests that specific pathways, potentially disrupted in severe COVID-19, are at play. Associations between delirium and changes in cardiac markers indexing parasympathetic activity have been reported, although not all markers show this relationship (Amit Patel et al., 2024). During severe COVID-19 infection, several mechanisms can affect vagal reflexes, such as hypoxia (Lemes and Zoccal, 2014), as a mechanism to maintain oxygen homeostasis. Furthermore, intubation, can trigger vagal reflexes leading to bradycardia (Jones et al., 2012). Additionally, inflammatory responses can also be mediated by or trigger posterior vagal activations, such as the case of cytokine storms leading to systemic organ failure (De Virgiliis and Di Giovanni, 2020). Indeed, a recent case-control study found that COVID ARDS was associated with marked bradycardia and severe HRV impairment, suggesting sympathovagal imbalance with vagal overtone as compared to non-COVID ARDS (Dumargne et al., 2024). Similar findings were made in another study comparing COVID-19 and all-cause sepsis (Kamaleswaran et al., 2021). Among COVID-19 patients, lower HRV indices associated with vagal modulation are generally associated with more severe disease (Pan et al., 2021) and mortality (Mol et al., 2021), consistent with our findings. While the exact pathways of the mechanisms involved in these interactions as well as their relations with the etiology and/or severity of the disease require more research, the pronounced effects on parasympathetic activity underscore the need for comprehensive monitoring of both sympathetic and parasympathetic functions.

### Limitations statement

The main limitations of our study include the relatively small sample size, potential confounding effects from sedation, and variability in patient severity and management, such as the use of medications like vasopressors that could affect brain or heart rhythms. Despite these limitations, the significant associations between brain-heart interactions and both mortality and acute encephalopathy remained after adjusting for sedation and non-neurological SOFA scores for mortality. Additionally, the absence of a non-COVID ARDS control group limits our ability to determine whether our findings are specific to COVID-19 compared to other causes of ARDS, whether septic or non-infectious.

## Conclusions

Our study demonstrates that bilateral brain-heart interactions in severe COVID-19 patients admitted to the ICU are compelling markers indicative of both mortality risk and acute encephalopathy in the ICU. These interactions evolve dynamically with the progression of the disease, with more pronounced dynamic changes correlating with milder conditions. Our findings highlight the importance of continuous monitoring of bilateral brain-heart interactions to understand these dynamics better and to develop tailored prognostic and therapeutic strategies based on the patients’ systemic conditions.

## Data Availability

All data produced in the present study are available upon reasonable request to the authors

## Abbreviations

ACE2: Angiotensin-converting enzyme 2
AE: Acute encephalopathy
ARDS: Acute respiratory distress syndrome
AUC: Area under the curve
CAM: Confusion Assessment Method
COVID-19: Coronavirus disease 2019
CPI: Cardiac parasympathetic index
CSI: Cardiac sympathetic index
ECG: Electrocardiogram
EEG: Electrocephalogram
FOUR: Full outline of unresponsiveness
HR: Heart rate
HRV: Heart rate variability
ICU: Intensive care unit
SARS-CoV-2: Severe acute respiratory syndrome coronavirus 2
SOFA: Sequential organ failure assessment
RASS: Richmond agitation sedation scale

## Author contributions

- Bertrand Hermann: Conceptualization, Data curation, Formal analysis, Funding acquisition, Investigation, Methodology, Project administration, Visualization, Writing – original draft, Writing – review & editing
- Sarah Benghanem: Conceptualization, Data curation, Formal analysis, Funding acquisition, Investigation, Writing – review & editing
- Estelle Pruvost-Robieux: Data curation, Writing – review & editing
- Tarek Sharshar: Conceptualization, Methodology, Supervision, Writing – review & editing
- Martine Gavaret: Data curation, Writing – review & editing
- Alain Cariou: Conceptualization, Investigation, Writing – review & editing
- Jean-Luc Diehl: Conceptualization, Investigation, Writing – review & editing
- Diego Candia-Rivera: Conceptualization, Formal analysis, Investigation, Methodology, Software, Supervision, Visualization, Writing – original draft, Writing – review & editing

