## Supplementary material for "Brain-heart interactions are associated with mortality and acute encephalopathy in ICU patients with severe COVID-19"

1) **Supplementary table 1. Univariable Cox models associations with cumulative rate of death and encephalopathy within 28 days of T1**

|  | **Death** | | **Encephalopathy** | |
| --- | --- | --- | --- | --- |
| **Independant Variables** | **HR [95% CI]** | **p-value** | **HR [95% CI]** | **p-value** |
| CSI → Brain | 0.21 [0.05-0.85] | 0.022 | 2.76 [1.18-6.42] | 0.016 |
| CPI → Brain | 0.24 [0.07-0.77] | 0.016 | 2.32 [1.09-4.93] | 0.025 |
| Brain → CSI | 1.66 [0.9-3.06] | 0.098 | 0.81 [0.57-1.15] | 0.24 |
| Brain → CPI | 2.11 [0.98-4.53] | 0.068 | 0.38 [0.18-0.8] | 0.005 |
| Non-neurological SOFA | 1.17 [0.94-1.46] | 0.147 | 0.85 [0.74-0.97] | 0.015 |
| RASS | 0.37 [0.12-1.13] | 0.037 | 1.34 [0.91-1.96] | 0.157 |
| Sedation rate (mg/10kg/h) | 1.61 [0.9-2.91] | 0.123 | 0.95 [0.62-1.44] | 0.804 |
| Analgesic rate (mg/10kg/h) | 1.19 [0.69-2.05] | 0.538 | 0.96 [0.66-1.41] | 0.839 |
| Sedation duration (days) | 0.99 [0.92-1.07] | 0.806 | 1.01 [0.97-1.05] | 0.687 |
| Sedative cumulative dose (mg/kg) | 1 [0.98-1.02] | 0.723 | 1.01 [0.99-1.02] | 0.434 |
| Analgesic cumulative dose (mg/kg) | 1 [0.99-1.01] | 0.937 | 1 [1-1.01] | 0.516 |

2) **Supplementary table 2. Multivariable Cox models adjusting on sedative/analgesic regimen.**

| **Mortality at D28 from T1** | | | | | | |
| --- | --- | --- | --- | --- | --- | --- |
| **Multivariate models** | **Sedative and Analgesic rate** | | **Sedative and Analgesic cumulative dose** | | **Sedative/Analgesic duration** | |
| **Independant Variables** | **aHR [95% CI]** | **p** | **aHR [95% CI]** | **p** | **aHR [95% CI]** | **p** |
| **CSI → Brain** | 0.16 [0.04-0.69] | **0.014** | 0.17 [0.04-0.69] | **0.013** | 0.2 [0.05-0.81] | **0.025** |
| **CPI → Brain** | 0.18 [0.06-0.6] | **0.005** | 0.22 [0.07-0.69] | **0.009** | 0.24 [0.07-0.78] | **0.017** |
| **Recovery from coma and delirium at day 28 from T1** | | | | | | |
| **Multivariate models** | **Sedative/Analgesic rate** | | **Sedative/Analgesic cumulative dose** | | **Sedative/Analgesic duration** | |
| **Independant Variables** | **aHR [95% CI]** | **p** | **aHR [95% CI]** | **p** | **aHR [95% CI]** | **p** |
| **Brain → CPI** | 0.38 [0.18-0.8] | **0.011** | 0.32 [0.14-0.73] | **0.007** | 0.3 [0.13-0.72] | **0.006** |
| **CSI → Brain** | 2.79 [1.2-6.52] | **0.018** | 3.27 [1.25-8.56] | **0.016** | 3.22 [1.23-8.48] | **0.018** |
| **CPI → Brain** | 2.55 [1.14-5.73] | **0.023** | 2.33 [1.04-5.26] | **0.041** | 2.31 [1.08-4.92] | **0.031** |

*Supplementary table 2. Multivariable Cox regression models with adjustment on sedative/analgesic.*

*Results from three separate models accounting for the potential effect of sedative/analgesic : left: adjustment on instantaneous sedative rate and analgesic rate at T1; center: adjustment on cumulative doses of sedative and analgesic at T1; right: adjustement on duration of sedative/analgesic prior to T1.*

3) **Supplementary table 3. Predictive performances of ECG, EEG and brainheart markers at T1 with ICU mortality delayed awakening**

| **Markers** | **Sen (%)** | **Spe (%)** | **PPV (%)** | **NPV (%)** | **AUC** |
| --- | --- | --- | --- | --- | --- |
| CSI | 50 [8-92] | 84 [32-100] | 50 [28-100] | 83 [77-94] | 0.56 [0.33-0.78] |
| CPI | 42 [8-92] | 92 [43-100] | 60 [29-100] | 83 [77-96] | 0.57 [0.35-0.79] |
| EEG broadband power | 75 [33-100] | 78 [43-100] | 55 [34-100] | 92 [82-100] | 0.75 [0.56-0.93] |
| CSI → Brain | 67 [42-100] | 97 [59-100] | 88 [40-100] | 90 [82-100] | 0.77 [0.59-0.96] |
| CPI → Brain | 75 [50-100] | 92 [78-100] | 75 [50-100] | 92 [84-100] | 0.79 [0.6-0.98] |
| Brain → CSI | 67 [33-100] | 86 [46-100] | 64 [35-100] | 89 [82-100] | 0.74 [0.55-0.93] |
| Brain → CPI | 58 [33-100] | 95 [41-100] | 78 [34-100] | 88 [82-100] | 0.74 [0.56-0.92] |

Sen : sensitivity ; Spe : specificity ; PPV : positive predictive value; NPV : negative predictive value; AUC : area under the ROC curve.

4) **Supplementary table 4. Predictive performances of ECG, EEG and brainheart markers at T1 with ICU mortality delayed awakening**

| **Markers** | **Sen (%)** | **Spe (%)** | **PPV (%)** | **NPV (%)** | **AUC** |
| --- | --- | --- | --- | --- | --- |
| CSI | 46 [12-69] | 86 [64-100] | 87 [73-100] | 45 [37-58] | 0.52 [0.34-0.7] |
| CPI | 35 [8-100] | 93 [14-100] | 86 [68-100] | 43 [37-100] | 0.5 [0.31-0.69] |
| EEG broadband power | 77 [38-100] | 64 [21-100] | 81 [70-100] | 64 [44-100] | 0.69 [0.51-0.87] |
| CSI → Brain | 58 [19-96] | 86 [29-100] | 85 [70-100] | 50 [39-83] | 0.61 [0.43-0.79] |
| CPI → Brain | 81 [15-100] | 57 [21-100] | 79 [69-100] | 58 [39-100] | 0.61 [0.42-0.8] |
| Brain → CSI | 65 [42-85] | 86 [57-100] | 88 [74-100] | 56 [43-73] | 0.62 [0.44-0.81] |
| Brain → CPI | 88 [73-100] | 86 [64-100] | 92 [81-100] | 79 [61-100] | 0.8 [0.63-0.97] |

Sen : sensitivity ; Spe : specificity ; PPV : positive predictive value; NPV : negative predictive value; AUC : area under the ROC curve.
